# Towards Improved Social Distancing Guidelines: Space and Time Dependence of Virus Transmission from Speech-driven Aerosol Transport Between Two Individuals

**DOI:** 10.1101/2020.08.31.20185439

**Authors:** Fan Yang, Amir A. Pahlavan, Simon Mendez, Manouk Abkarian, Howard A. Stone

## Abstract

It is now recognized that aerosol transport contributes to the transmission of the SARS-CoV-2 virus. Here we improve existing social distancing guidelines for airborne pathogens, which are typically given in terms of distance with vague statements (if any) about contact times. Also, estimates of inhalation of virus in a contaminated space usually assume a well-mixed environment, which is realistic for some, but not all, situations. In particular, we consider a local casual interaction of an infected individual and a susceptible individual, both maskless, account for the air flow and aerosol transport characteristics of speaking and breathing, and propose social distancing guidelines that involve both space and contact time, based on a conservative model of the interactions.

## I. INTRODUCTION

Transmission of the virus SARS-CoV-2 during the presymptomatic and asymptomatic stages of the disease COVID-19 is estimated to be responsible for more than half of all of the cases [1, 2]. The recognition that aerosols play a significant role in the transmission of SARS-CoV-2 raises the issue of quantifying the air exchange between asymptomatic individuals, who are sources of the virus, and healthy individuals, who are susceptible to infection. A common model for characterizing this situation is to evaluate the probability of infection based on the number *N* of virions inhaled, relative to a characteristic dose *N*_*inf*_ that, on average, produces infection; the corresponding probability of infection *p* is then estimated as [3]

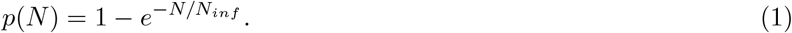

Past studies of the transmission of other viruses provide insights into the risk of infection by performing room-scale averages of the virion concentration, e.g. [4, 5]. Recent studies applied to SARS-CoV-2 have similar features and allow an estimate of the number of inhaled virions by a susceptible individual in a space, e.g., a bathroom, airplane, laboratory, etc. [6–8]. Such well-mixed models are appropriate when the asymptomatic source individuals are well removed from healthy individuals, so that the time scale to mix in the environment is faster than the time for direct exchange of air between a source and a receiver.

Here, we are concerned with casual conversations, where *local* air exchange between individuals is the dominant factor in determining the infection probability. In contrast with violent expiratory events, such as coughing and sneezing, which have been shown to be an important pathway in transmission of pathogens from symptomatic individuals [9, 10], we focus on speaking and breathing, which are relevant to transmission during presymptomatic and asymptomatic stages. Some earlier studies have characterized qualitative features of the respiratory flow between two people using numerical simulations and experiments with mannequins [11, 12] and a recent study has numerically probed spatial features of drop and aerosol transport between two people where one is an infected speaker [13]; also, see the reviews [14–16].

We consider a poorly ventilated environment, in which the separation distance between an asymptomatic speaker and a healthy interlocutor is comparable to those recommended by the World Health Organization (1 m) and the Center for Disease Control (CDC) in the United States (2 m). Indeed, we can imagine that such local, air flow-driven virus exchange may occur in conversations at parties, across the table at lunch or in a conference room, on a train, etc. Recent studies provide estimates of the characteristic infection dose, *N*_*inf*_ ≈ 100 − 1000, and, consistent with other modeling efforts, below we take the conservative value *N*_*inf*_ = 100 [6, 7, 17]. We assume the transport of aerosolized virions is that of a passive scalar, as justified below, and use a model of the time and space dependence of air flow in speech from our recent work [18]. Thus, to characterize the risk of infection we provide a space-time diagram of social distancing. These assessments, in the absence of mask wearing, should be combined with other *global* estimates to better understand the possible dynamics of pathogen exchange in different situations.

## II. TRANSPORT FEATURES OF SPEECH

### II.1. Assumptions

Given the complexity of the speech-driven aerosol transport, we make the following simplifying assumptions in our analysis: (1) Consistent with the estimates of the fluid dynamics below, and recently published experiments and numerical simulations [18], the expiratory flow from speech is a conical, quasi-steady jet, with a horizontal extent set by the speaking time. (2) Both individuals are maskless and in a poorly ventilated environment, which we take as those spaces where the ambient flow speed is *O*(1) cm/s. (3) The infected individual speaks continuously and the susceptible individual only listens (and breathes). The disturbance imparted by the susceptible receiver on the on-coming jet is neglected; this assumption will be justified in section II.2. (4) The ambient relative humidity (RH), which is the ratio of partial pressure of water vapor to the equilibrium saturation vapor pressure of water, is not too large, so that droplets from speech will evaporate quickly and form aerosols. In section II.3 we provide an example for RH=50%.

We note that these assumptions yield a conservative estimate for the worst-case scenario for pathogen transmission in a local casual interaction. These assumptions also apply to a susceptible individual that joins a group and stands opposite an asymptomatic person who has been speaking for some time.

### II.2. Conical, quasi-steady air flows characterize exhalation during speech

The dynamics of the air flows in speaking and breathing are controlled by the Reynolds number, Re = 2*av*_0_*/ν*, where *a* is a typical length of the opening of the mouth, *v*_0_ is the average speed of the flow at the mouth (typical values are given in Tables I and II), and the kinematic viscosity of air *ν* ≈ 1.5 × 10^*−*5^ m^2^/s. Thus, the Reynolds numbers are relatively high (*Re* ≈ 100 − 1000). Utilizing laboratory experiments of the air flow during speaking, numerical simulations of the Navier-Stokes equations for pressure-driven flows from an orifice, which mimics speech, and a mathematical model of these dynamics, our recent work [18] has shown that maskless speaking and breathing in poorly ventilated environments can produce a conical quasi-steady, turbulent jet after a few seconds, as shown in Fig. 1. Sufficiently far away from the mouth, the unsteadiness of the inhaling/exhaling signal is barely visible and the jet behaves similar to a turbulent jet with constant flow rate. A typical laboratory measurement of the flow field produced by breathing is displayed in Fig. 1(a), while a simulation of the air flow produced by speaking is shown in Fig. 1(b). The typical cone half angle *α* is about 10 – 14°. Hence, the area of the cone will envelop the size of a human head already at 50 cm separation distance. To illustrate the magnitude of exhaled concentrations relevant to typical social distances, in the simulation shown in Fig. 1(b) we highlight a concentration contour *c* = 0.05 (with *c* = 1 at the mouth), which is comparable to the scale of the head a distance 1.6 m away (the right limit of the figure). Hence, non-negligible exhaled concentrations are found at distances comparable to social interactions.

**TABLE I:** Typical values related to speaking reported in the literature.

|  |  |
| --- | --- |
| SARS-CoV-2 concentration in saliva $c_v$ | average: $7 \times 10^6 \text{ mL}^{-1}$, maximum: $2 \times 10^9 \text{ mL}^{-1}$ [31] |
| total droplet production rate $j_0$ | 140 – 770 nL/min [34] |
| flow velocity at the mouth $v_0$ | 1 – 5 m/s [20, 35–39] |
| inhalation volume flux $Q_r$ | 0.1 L/s [6] |
| half cone angle $\alpha$ | 10 – 14° [18] |

**TABLE II:** Parameters used in Figs. 3 and 4 for characterizing SARS-CoV-2 viral contamination by an asymptomatic individual speaking in a poorly ventilated space.

| $\phi_0$ | $c_v$ | $N_{inf}$ | $Q_r$ | $a$ | $\alpha$ |
| --- | --- | --- | --- | --- | --- |
| $6 \times 10^{-9}, 1 \times 10^{-8}$ (only in Fig. 4 (b)) | $7 \times 10^6 \text{ mL}^{-1}$ | 100 | 0.1 L/s | 1 cm | 12° |

**FIG. 1:**
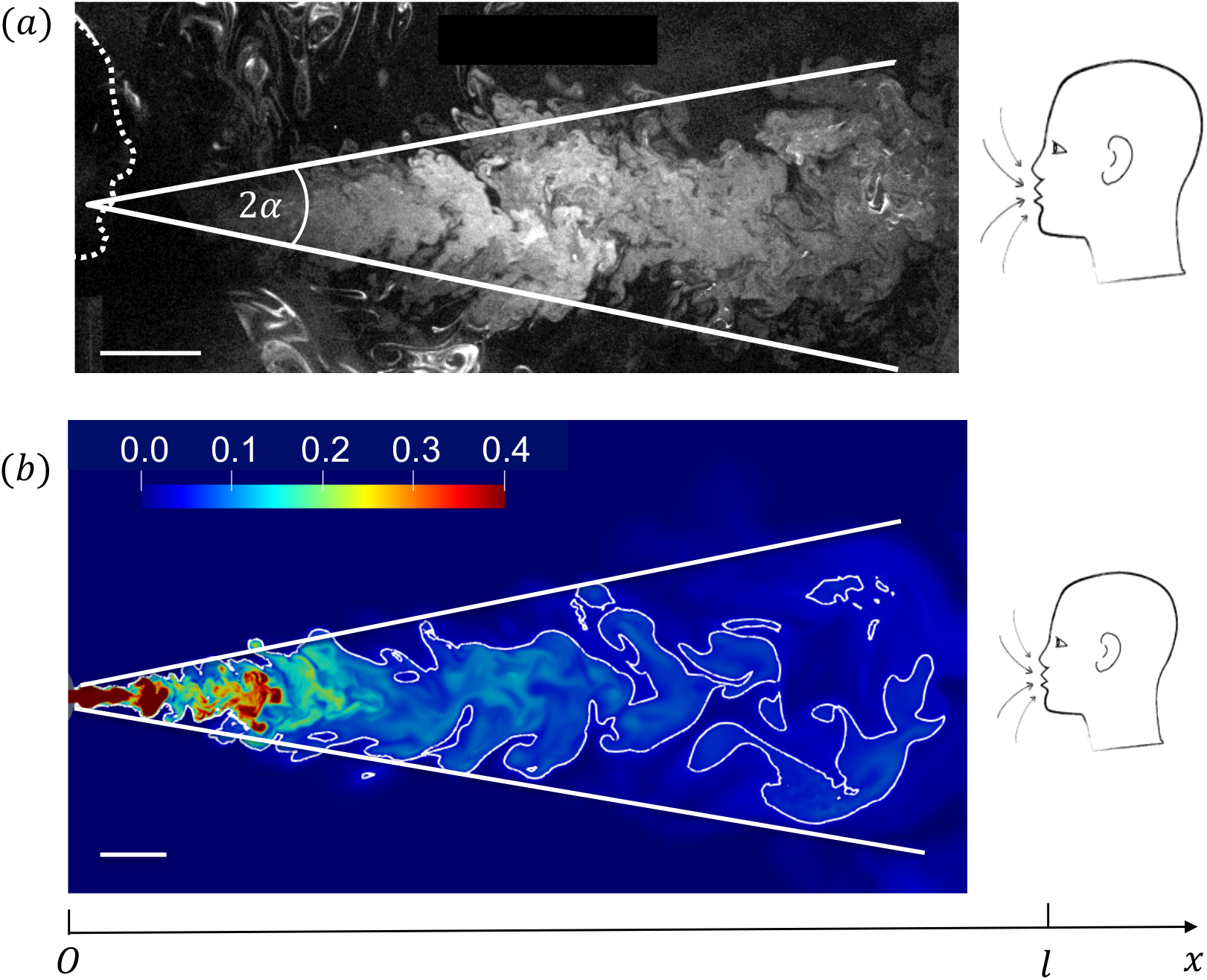
Conical flows from breathing and speech. (a) Flow visualization of the conical jet produced by breathing from the mouth, adapted from Ref. [18]. Speaking can produce similar conical jets. The cone angle is 2*α*. A (susceptible) receiver, shown with a head sketched on the right, is facing the asymptomatic (infected) breather/speaker at a distance *ℓ*. (b) Simulated transport of exhaled material during repeated speech. Results of numerical simulations from Ref. [18]: example of the instantaneous field of a passive scalar used to visualize the dilution of the air exhaled from an infected speaker, displayed over a vertical symmetry plane after 9.5 cycles of periodic speaking-inhaling (each cycle lasts 4 s), with a volume per breath of 0.75 L. The color map shows the concentration field *c* of the passive scalar with *c* = 1.0 at the exit of the mouth and *c* = 0 in the ambient far-field (the scale is saturated here to better visualize the field close to the receiver). An iso-concentration line at *c* = 0.05 is displayed to help quantify the dilution levels far from the mouth of the speaker to the left. Both scale bars are 10 cm.

We note that our experiments show that during speaking the effect of buoyancy at the scale of a few meters of jet motion (Fig. 1(a)) does not cause a substantial vertical motion beyond the size of a human head. Moreover, due to entrainment of the surrounding air, the thermal signature will decay inversely with distance, comparable to a passive scalar such as an aerosol, as discussed below, which further diminishes the buoyancy effects.

As illustrated in Fig. 1, for the typical Reynolds numbers *Re* ≈ 100 − 1000, the expiratory flow is approximately jet-like and entrains surrounding air, whereas we expect the inhalatory flow to be approximately hemi-spherical, as sketched in Fig. 2; see [18, 19]. As a consequence of the asymmetry of inhalation and exhalation, only a small portion of the inhaled breath comes from the exhaled material of the same individual. This may be verified easily by a simple model: if we assume the volume of air inhaled and exhaled in one breathing cycle is the same, denoted by *V*, then the radius *R* of the inhaled hemisphere is *R* = (3*V/*2*π*)^1*/*3^. In the volume to inhale, only a small conical portion (the shaded region in Fig. 2) is occupied by the exhaled material, whose volume is *V*_*e*_ ≈ *π*[(*R*+*a* cot *α*)^3^ tan^2^ *α* − *a*^3^ cot *α*]*/*3. Using *V* = 0.5 L [20] and *α* = 12°, we have *V*_*e*_*/V* ≈ 0.1. Indeed, exploiting the series of numerical simulations of periodic breathing and speaking published in Ref. [18], which used an elliptic mouth with minor and major axes respectively 1 cm and 1.5 cm, and a fixed cycle duration of 4 s, then for very different flow rate signals, and a volume per breath between 0.5 L and 1.0 L (with equal volumes exhaled and inhaled), we confirmed that typically 10% of the exhaled material from one breath was inhaled in the next inspiration. Systematic experiments mimicking human breathing show that the re-inhalation ratio is 2–10% for *Re* = 100 − 1000 [19]. This means that when a person inhales, they mainly inhale the ambient air. It is thus the environment around the head of the (healthy) person that needs to be characterized to develop an estimate of the risk of pathogen uptake in these close encounters.

**FIG. 2:**
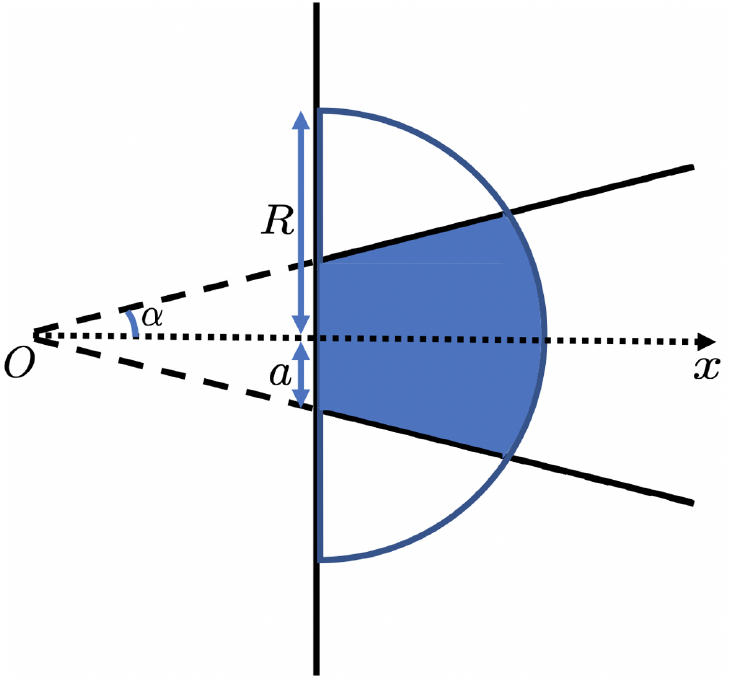
Sketch of ideal respiratory flow regions. The expiratory flow region is a cone with half-angle *α* and apparent origin at *O*. The mouth is modelled as a circle of radius *a*, located at *x* = *a* cot *α*. The inhalatory flow region is a hemisphere with radius *R* (determined by the inhaled volume, see text) and centered at *x* = *a* cot *α* (the exit of the mouth). In the inhaled volume, only the shaded region is from the previous exhalation.

### II.3. Evaporating drops and aerosols as passive tracers following the air flow

We next comment about time scales. Experiments show that it takes 30 to 50 seconds for the jet to reach a distance 3 m for steady speaking [18]; the jet can reach 2 m in even 20 s. The radii of droplets produced by speaking are found to be typically around or smaller than 5 *µ*m [6, 21, 22]; drops of smaller sizes are traditionally categorized as aerosols. It takes about 0.1 s for a water droplet of radius 5 *µ*m (and about 2 s for radius 20 *µ*m) to evaporate at RH=50% [23], so most small droplets produced by speaking should evaporate rapidly to become aerosol particles at the beginning of the jet spreading. We note that a recent numerical study shows that the the lifetime of droplets is significantly extended in a puff mimicking a strong cough, mainly due to the high local RH(=100%) and the flow field in the puff [24].

During Δ*t* = 50 s, an aerosol particle of radius *r*_*p*_ = 1 *µ*m and density *ρ*_*p*_ = 10^3^ kg/m^3^ sediments a distance of 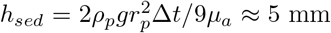, where *g* = 9.8 m^2^/s is the gravitational acceleration and *µ*_*a*_ = 1.8 × 10^*−*5^ Pa·s is the viscosity of air. Therefore, at the scale of a social interaction, we can ignore sedimentation of the small particles. Moreover, the Stokes number, 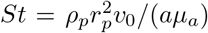, which characterizes the tendency of the more dense particles to deviate from the streamlines of the air flow, has a typical magnitude *St* = *O* 10^*−*2^ ≪ 1. So, we treat the aerosol particles as passive tracers that follow the jet flow.

## III. AN ESTIMATE OF RISK FROM VIRUS INHALATION

When an asymptomatic subject exhales or speaks, small droplets carrying virus can spread to a receiving interlocutor. To characterize the important aspects, we note that the drop size distribution [6, 21, 22, 25–27], the influence of loudness and phonetic features on droplet production rates [28–30], and viral densities in saliva [31] have been reported in the literature. Here, we use these measurements, together with the flow field characteristics sketched in Section II.2, to quantify the amount of virus that will reach the receiver in a poorly ventilated space, and so provide a measure of the risk of infection. The model allows quantitative assessment of the impact of separation distance and time of interaction.

The speaker’s mouth can be approximated as a circle with radius *a*. Denoting the cone angle as 2*α* and the spatial axis from the cone vertex as *x* (see Fig. 1), the cross-sectional area of the jet beyond the mouth is *A*(*x*) = *π*(*x* tan *α*)^2^. Note that the size of the turbulent jet increases as it propagates due to entrainment of the surrounding air [9, 18, 32]. In a steady jet, with average velocity *v*(*x*), the momentum flux, which is proportional to *v*^2^(*x*)*A*(*x*), is constant. Therefore *v*(*x*) = *v*_0_*a/x* tan *α*, where *v*_0_ is the flow velocity at the mouth. Denoting the virus concentration in the saliva of the asymptomatic speaker as *c*_*v*_ (number virions/volume saliva) and the volume fraction of droplets at the mouth as *ϕ*_0_ (volume droplets/volume air), then the total droplet volume production rate in speaking is *j*_0_ = *πa*^2^*v*_0_*ϕ*_0_ (volume droplets/time). It follows that the total emission rate of virus is *I*_0_ = *c*_*v*_*j*_0_ (number virions/time). Within a quasi-steady approximation [18], and denoting *ϕ*(*x*) as the volume fraction of droplets in the jet, the flux of virus in the steady jet *c*_*v*_*ϕ*(*x*)*v*(*x*)*A*(*x*) is also constant and equal to *I*_0_. Thus, we conclude *ϕ*(*x*) = *ϕ*_0_*a/x* tan *α*. The variation with distance as *x*^*−*1^ of the pathogen concentration (*c*_*v*_*ϕ*(*x*)) is characteristic of speaking.

We now consider a susceptible individual at a distance *ℓ* in front of an infected speaker (Fig. 1). Assuming the average inhalation volume flux of the receiver (at *x* = *ℓ*) to be *Q*_*r*_, then the intake dose of the receiver is

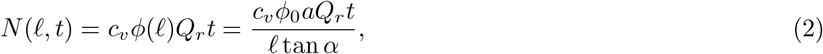

over a period of time *t*. This result does not depend explicitly on the speed of the air flow produced by the speaker. Also, we note that as the droplets evaporate *c*_*v*_ will increase and *ϕ*(*x*) will decrease, however, the virus concentration *c*_*v*_*ϕ*(*x*) (number virions/volume air) is unaffected by evaporation.

Typical values of the variables in equation (2) are summarized in Table I. The typical range of *ϕ*_0_ is calculated from Table I to be 2 × 10^*−*9^ − 1 × 10^*−*8^. Note that the virus emission rate *I*_0_ in breathing has been measured by collecting respiratory droplets and aerosols in 30 min from infected people [33]. The average value is around 300 min^*−*1^, which is smaller than *I*_0_ in speaking: for an average infected patient (*c*_*v*_ = 7 × 10^6^ mL^*−*1^, *v*_0_ = 3 m/s), *I*_0_ ≈ 1 × 10^3^ − 5 × 10^3^ min^*−*1^.

Although the typical “infectious dose” of SARS-CoV-2 is unknown, estimates have been provided. Consistent with other studies, we approximate *N*_*inf*_ conservatively as *N*_*inf*_ = 100 [6, 7]. Based on this criterion and equation (2), we can plot the probability of infection *p* versus distance *ℓ* and time *t* as shown in Fig. 3. The results show the importance of distancing and decreasing contact time for reducing the probability of infection. In order to use a simple terminology we will define the boundary between lower- and higher-risk situations with the probability *p*(*N* = *N*_*inf*_) = 1 − *e*^*−*1^ = 0.63. From Fig. 3, a distance of *ℓ* = 1 m can maintain a lower risk for an interaction of only 8 min; for *ℓ* = 2 m, the interaction time should be restricted to less than 16 min. If one wants to maintain the probability of infection under 0.2, the corresponding contact times should be less than 2 min and 4 min respectively, as shown in Fig. 3. We remind the reader that these estimates are based on a single maskless infected individual in a poorly ventilated space (using average virus parameters reported in the literature).

**FIG. 3:**
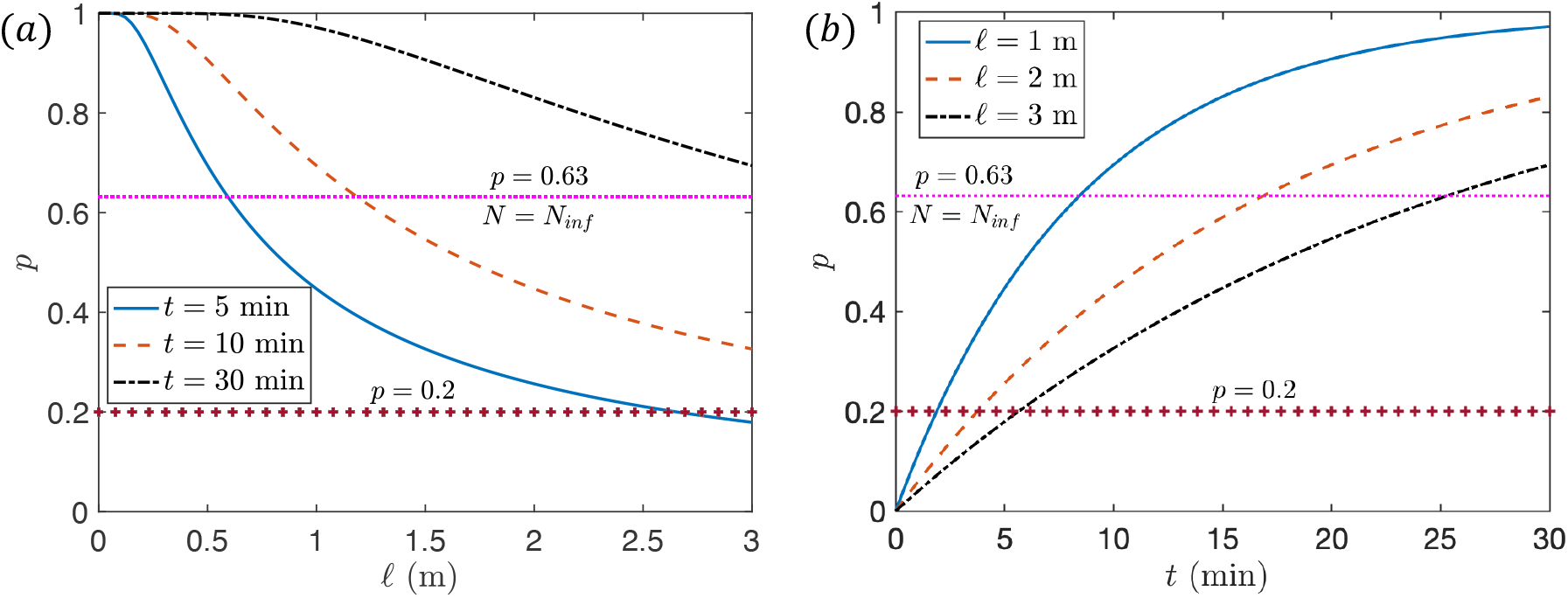
Dependence of infection probability *p* on (a) distance *ℓ* and (b) speaking time *t* (for interaction with a single maskless asymptomatic speaker). Values of *ℓ* or *t* for each curve are given in the legends. The spatial distance is truncated at 3 m, beyond which we expect the ambient flows to become important even in a poorly ventilated space [18]. Other parameters are listed in Table II. Two horizontal lines *p* = 0.63 (or equivalently *N* = *N*_*inf*_) and *p* = 0.2 are plotted.

Next, we present a space-time, social distancing diagram of infection risk. In Fig. 4(a), we illustrate the probability of infection (yellow is high and blue is low), with spatial separation indicated on the horizontal axis and contact time on the vertical axis. This figure makes clear that at distances typically discussed for social distancing, longer contact times in speaking engagements with an asymptomatic individual increase the probability of infection.

**FIG. 4:**
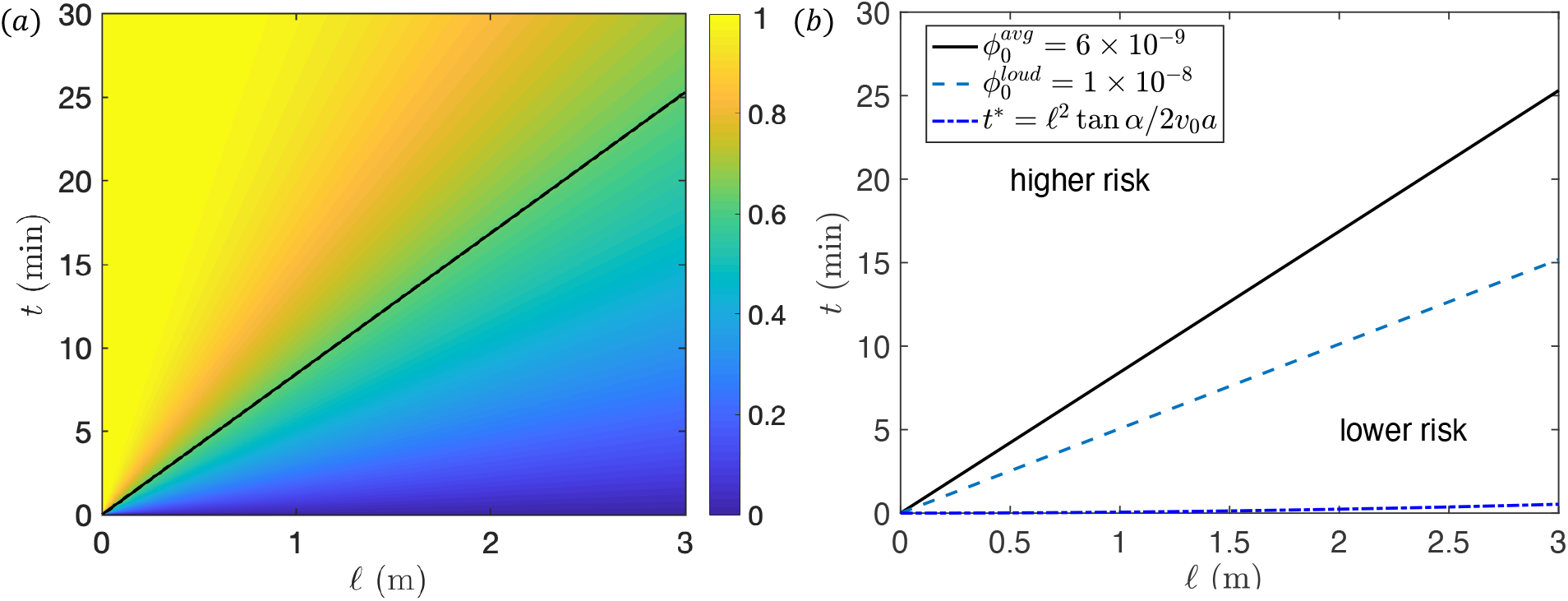
Space-time diagram of infection risks. The color map (a) shows the infection risk *p* as a function of distance *ℓ* and contact time *t* with *ϕ*_0_ = 6 × 10^*−*9^, in which the straight line indicates *p* = 0.63. (b) shows the effects of *ϕ*_0_. The infection risk is considered lower (higher) in the region below (above) the straight lines, where *p* = 0.63 or equivalently, *N* = *N*_*inf*_ is proposed as the boundary for higher and lower risks. In (b) *t*\* indicates the time for the jet to reach *ℓ* in the initial spreading stage. Below *t*\* is the no-risk region. The plot is for a susceptible individual interacting with a single maskless asymptomatic speaker in a poorly ventilated space.

Experiments show that the production rates of droplets increase with the loudness of speech [28–30], which is approximately examined in Fig. 4(b) by using 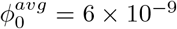 (typical speech) and 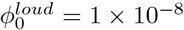 (loud speech). We observe that for *ℓ* = 2 m and 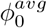, and based on *N* = *N*_*inf*_ as the estimate for increased risk, it is higher risk to speak with an asymptomatic individual for more than 16 min, but with loud speech 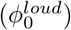 for more than 10 min. According to this model, when talking over 25 min, there is a higher risk of infection even at a separation distance of 3 m.

We note that for a speech-driven flow, the time for the jet to reach a distance *ℓ* is *t*\*(*ℓ*) = *ℓ*^2^ tan *α/*2*v*_0_*a* [18]. In Fig. 4(b), the region below *t*\* is therefore a “no-risk” region since there has not been time for any exhaled material to reach the receiver. Experiments show that *t*\*≈ 30 − 50 s for the jet to reach *ℓ* = 3 m [18], which matches the calculated result *t*\* = 32 s using parameters in Table II. The value of *t*\* is usually much smaller than the critical time *t* separating the higher- and lower-risk regions in Fig. 4(b).

In all of above discussions an average virus concentration in saliva *c*_*v*_ = 7 × 10^6^ mL^*−*1^ is assumed. However, for a patient with a very high viral load in the saliva, *c*_*v*_ can be as high as 2 × 10^9^ mL^*−*1^, which is 300 times larger than the average value [31]. Consequently, the time *t*_*s*_ for *N* (*ℓ, t*_*s*_) = *N*_*inf*_ at *ℓ* = 3 m is *t*_*s*_ ≈ 5 s (using 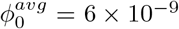 and other parameters listed in Table II), which is much smaller than *t*\* and means that the virion uptake by the receiver will surpass *N*_*inf*_ almost as soon as the jet reaches them. Therefore, the infection probability for speaking with a patient with a very high viral load in the saliva (a potential superspreader) even for less than 1 min is high at a 3 m separation. Wearing masks can block the formation of jet, filter droplets and aerosols, and thus mitigate against the transmission of airborne pathogens [40].

### IV. CONCLUDING REMARKS

We analyzed the spatial and temporal dependence of local virus transmission between a speaking asymptomatic individual and a susceptible listening individual in the absence of mask wearing and in a poorly ventilated environment. We used recent quantitative characteristics of speech [18] and typical viral loads of COVID-19 infected individuals. We show that both social distancing and decreasing contact time are important to keep the probability of infection low. Our analysis suggests that the mask-free social distancing guidelines, 1 m according to WHO and 2 m according to CDC in the United States, should be accompanied by contact-time guidelines. In particular, our estimates above, using typical values reported in the literature for droplet production rates and viral loads in saliva, suggest that probability of infection is relatively high for ≈ 8 min of contact time for 1 m separation distance and ≈ 16 min for 2 m of separation distance. If the infected speaker has a very high viral load in the saliva, the infection risk is high within less than 1 min for *ℓ* = 3 m separation.

Also, we emphasize that the model presented here is for the worst-case scenario when the infected individual is actively speaking and the susceptible (maskless) individual is a passive listener. Future research is required for a better understanding of the more complex flow between two (or more) speakers and its impact on aerosol transmission. Similar estimates and guidelines for the importance of contact time have been proposed recently for ventilated spaces, based on a well-mixed room model [41]. The important point to emphasize is that in social situations there are also time restrictions that should be imposed to lower the risk of infection.

## Data Availability

All data are available and provided in the manuscript.

## ACKNOWLEDGMENTS

We thank the National Science Foundation for support via the RAPID grant CBET 2029370 (Program Manager is Ron Joslin). We also thank the IRN “Physics of Living Systems” (CNRS/INSERM) for travel support for M.A.

